# A Tale of Two Lenses: Emergency department indoor-air hybrid-capture metagenomics complements wastewater by adding a human-focused respiratory virus perspective

**DOI:** 10.64898/2026.03.13.26348311

**Authors:** Mustafa Karatas, Sarah Gorissen, Jens Swinnen, Caspar Geenen, Katrien Van Dyck, Lize Cuypers, Bieke Tack, Edouard Hosten, Mandy Bloemen, Elke Wollants, Beau Verschueren, Lies Laenen, Kurt Beuselinck, Annette Schuermans, Marc Van Ranst, Marc Sabbe, Jelle Matthijnssens, Emmanuel André

## Abstract

**Background:** Continuous, non-invasive viral surveillance is essential to monitor emerging pathogens and guide public health responses. Most environmental surveillance studies use targeted qPCR approaches, and comparisons between wastewater and indoor air surveillance remain limited. We aimed to compare the utility of emergency department indoor air and urban wastewater for tracking circulating viruses and resolving genomic information.

**Methods:** We conducted a matched-pair study comparing 19 weekly indoor air samples from the central ventilation exhaust shaft of an emergency department and 19 24-hour composite municipal wastewater samples in Leuven, Belgium, from December 2024 to April 2025. Both sample sets were processed using probe-based hybrid-capture viral metagenomics targeting over 3000 viral species, using influenza A as a clinically relevant test case.

**Findings:** Wastewater captured higher overall viral diversity (233 versus 106 species) and more complete genomes compared to indoor air, showing a relatively stable composition, mainly of enteric and animal-associated viruses. Indoor air demonstrated lower overall diversity but was enriched for respiratory viruses, including influenza A, coronaviruses, metapneumovirus, and respiratory syncytial virus, and more frequently achieved high genome coverage for these pathogens. Although both sample types permitted influenza A subtype characterization, influenza A genomes from wastewater were often less well covered. When coverage thresholds were met, indoor air supported targeted antiviral resistance-site screening for influenza A and RSV-A.

**Interpretation:** Wastewater and indoor air generate distinct but complementary viromes. Wastewater acts as a diverse, population-level monitor for One-Health applications, whereas indoor air serves as a targeted, human-centric sentinel system facilitating further genomic characterization for respiratory viruses.

**Funding:** Mustafa Karatas is supported by a Research Foundation Flanders (FWO) fundamental research scholarship (number: 11P7I24N). C.G., L.C., E.H., S.G. and E.A. acknowledge support from the DURABLE project. The DURABLE project has been co-funded by the European Union, under the EU4Health Programme (EU4H), project no. 101102733.

**Research in context:** 

**Evidence before this study:** We searched PubMed for studies published between Jan 2000 and March 2024 using the terms “wastewater surveillance”, “metagenomics”, “indoor air”, and “viral metagenomics”. Previous studies have shown that wastewater surveillance can detect population-level viral circulation, and more recent work has explored indoor air sampling as a method for monitoring respiratory virus transmission. However, environmental metagenomic studies have largely examined these two sample types separately. Furthermore, most studies relied on untargeted sequencing approaches, which often yield fragmented genomes in these environments. To date, no study has systematically compared indoor air and wastewater using a comprehensive hybrid-capture viral metagenomics approach for virus surveillance.

**Added value of this study:** We conducted a matched comparison of indoor air from a hospital emergency department and municipal wastewater collected during the same weeks in Leuven, Belgium. We analyzed both sample types using an identical hybrid-capture viral metagenomics workflow targeting more than 3000 viral species. This design enabled a direct evaluation of how the two environmental surveillance lenses differ in viral diversity, genomic recovery, and epidemiological relevance. Wastewater captured broader viral diversity and a stable background dominated by enteric and animal-associated viruses, whereas indoor air captured more respiratory viruses and more frequently yielded high genome completeness for these pathogens. When genome coverage thresholds were met, indoor air data enabled influenza subtype identification and screening for antiviral resistance markers.

**Implications of all the available evidence:** Our findings support a layered environmental surveillance strategy in which different environmental samples provide complementary information. Wastewater offers a stable, population-level view of viral circulation and captures broad viral diversity, including human and animal-associated viruses. Indoor air sampling in human-dominated settings provides a more direct signal of respiratory virus circulation and can yield genomes suitable for subtype and mutation-level characterization. Combining these approaches could strengthen metagenomic surveillance frameworks by improving the interpretation of environmental viral signals, supporting early detection of emerging pathogens, and helping distinguish human virus circulation from environmental or animal-derived detections.

## Introduction

The COVID-19 pandemic and ongoing recent resurgence and geographic expansion of highly pathogenic avian influenza highlight the increasing frequency and impact of viral emergence and re-emergence.^1^ In a context of continuous viral evolution and virus- or strain-specific medical countermeasures, surveillance systems remain largely pathogen-specific and compartmentalized. Multiplex qPCR panels enable monitoring of predefined viruses but are inherently limited to a finite set of targets and provide little genomic resolution for subtype, variant, or mutation characterization^2^ Viral metagenomics expands surveillance beyond predefined targets and enables genome-resolved detection of both expected and unexpected viruses. Probe-based hybrid capture can further enhance sensitivity and genome recovery in low-biomass and high-background environments such as wastewater and indoor air^3–5^, facilitating in-depth genomic analyses. In return, genomics can provide direct information about the evolution of pathogens and potentially inform public health decisions.

Wastewater surveillance can provide broad population coverage and allow tracking epidemics in a community, but it is a composite sample, shaped by rainfall-related dilution and flow variation, and virus-specific shedding routes (stool/urine versus primarily respiratory shedding).^6,7^ In addition, viral RNA detected in wastewater may also include environmental or animal contributions depending on local infrastructure and urban ecology. Recent reports of highly pathogenic avian influenza (HPAI) signatures in wastewater illustrate this challenge of wastewater-based detections, ^8,9^ as they reflected animal or environmental sources and do not always indicate human infection. ^10^

Indoor air sampling is increasingly explored for respiratory pathogen surveillance because it can provide location-specific signals^11^ and can be more directly linked to human shedding, especially in built indoor environments such as hospitals^3^ or daycare centers^12,13^. In human-dominated settings, indoor air may provide a more human-centric view of circulating respiratory viruses than wastewater, but systematic, genome-resolved comparisons of wastewater and indoor air remain scarce.^14^

Here, we leveraged a comprehensive hybrid-capture viral metagenomics tool targeting more than 3,000 human- and animal-infecting viral species to conduct a matched comparison of University Hospitals Leuven emergency department indoor air and municipal wastewater in Leuven, Belgium, during the 2024–2025 influenza season. We quantified viral diversity, genome completeness, and community structure across both sample types (i.e. indoor air and wastewater) using a harmonized workflow for wet-lab and bioinformatics analyses. As a clinically relevant test case, we evaluated influenza A, a rapidly evolving respiratory virus for which subtype and mutation-level resolution are epidemiologically meaningful. Moreover, we verified whether indoor air genome recovery from viral metagenomics could support resistance-site screening for influenza A and RSV in case of sufficient genome coverage.

## Results

We conducted a matched-pair comparison study analyzing 38 environmental samples collected between December 2024 and April 2025 from Leuven, Belgium: 19 indoor air samples from the emergency department at University Hospitals Leuven and 19 corresponding wastewater samples from the municipal treatment plant of Leuven. Wastewater samples represent a city-level, 24-hour composite sample, whereas indoor air samples collected from the HVAC system reflect a hospital emergency department microenvironment aggregated over the course of one week. Hybrid-capture viral metagenomics was applied to all samples and compared with influenza A clinical data from University Hospitals Leuven.^15^

### 1. Wastewater identifies more virus species and more complete genomes overall

Across 19 weeks, wastewater yielded 233 viral species compared to 106 viral species in indoor air (Figure 1A). This disparity was largely driven by non-enveloped DNA viruses, with wastewater showing a substantially higher number of species for *Papillomaviridae* (61 vs. 35 species), *Parvoviridae* (45 vs. 13), *Circoviridae* (24 vs. 2), and *Anelloviridae* (21 vs. 1). Conversely, species counts for enveloped virus families were comparable or slightly higher in indoor air (*Coronaviridae*: 4 vs. 5; *Herpesviridae*: 5 vs. 6) (Figure 1B). Temporal analysis revealed distinct stability profiles: wastewater maintained a highly stable composition dominated by members of the families *Polyomaviridae, Parvoviridae* and *Circoviridae*, whereas indoor air composition showed stronger fluctuations due to episodic pulses of viruses such as herpesviruses (i.e., CMV, HSV-1), influenza A (*Orthomyxoviridae*) and rotavirus A (*Sedoreoviridae*, Figure 1C). Moreover, wastewater yielded higher genome recovery, with a median of 84 detections per sample with >75% completeness, nearly threefold higher than the median of 29 detections in indoor air (Figure 1D).

**Figure 1.**
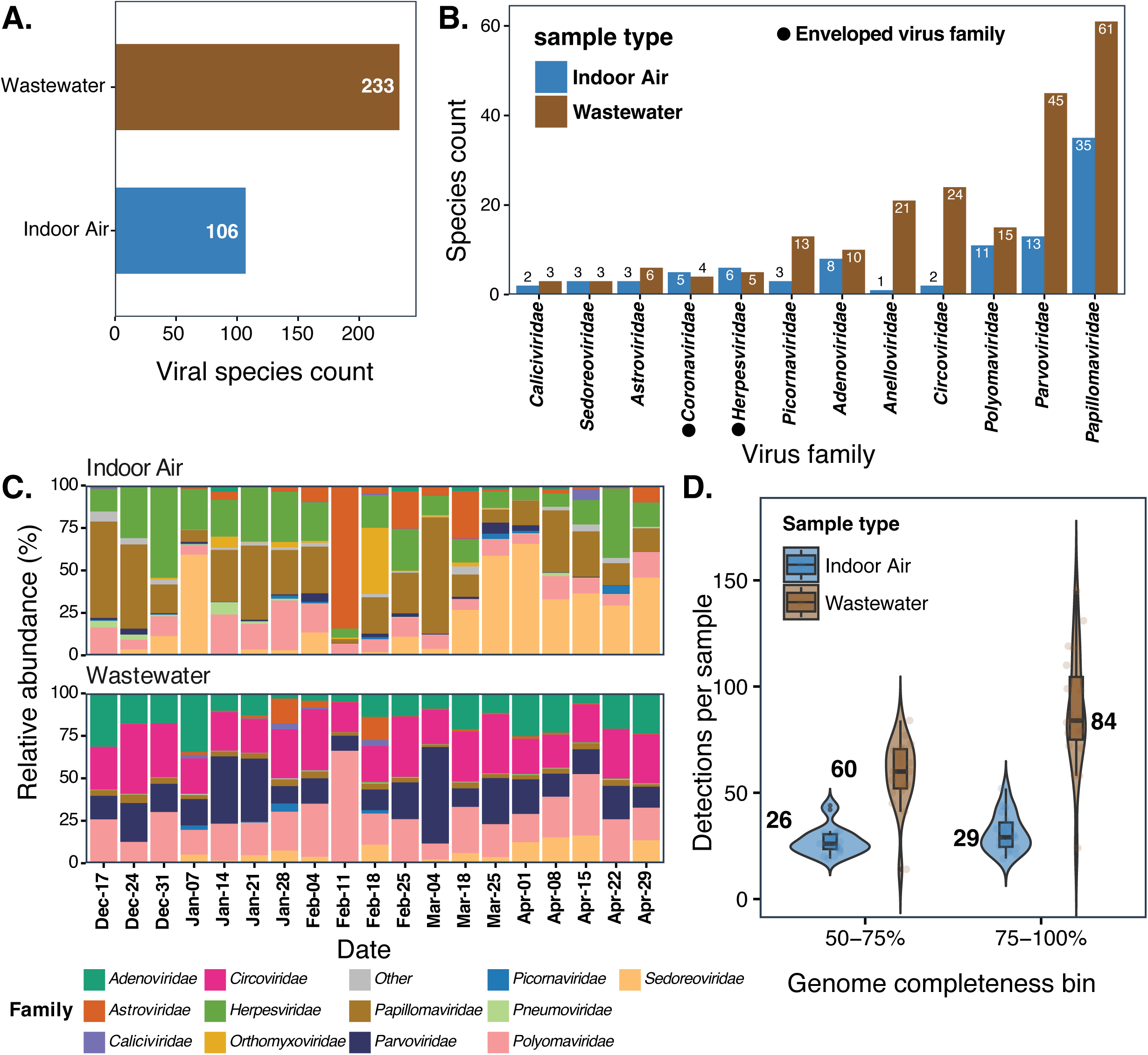
Comparative overview of viral community diversity and genomic information recovery from indoor air and wastewater surveillance. **A.** Total virus species detected across matched sample pairs (n=19 pairs; 38 total samples) by sample type. **B.** Number of species detected within the 12 most commonly identified viral families. **C.** Temporal family-level relative abundance (% of total reads per sample) across matched sampling dates, stratified by sample type. **D.** Distribution of viral detection counts per sample, stratified by genome completeness bins (≥50-75% and ≥75-100% coverage of reference sequences). Violin plots show distribution shapes overlaid with box plots (quartiles and median) and individual sample points.

### 2. Indoor air is more respiratory- and human-specific, wastewater is more stable and captures more diversity, including animal-associated viruses

We next compared sample type-specific patterns for selected pathogenic viruses using genome completeness as the primary measure of recovered genomic information. Respiratory viruses, including influenza A and B, and members of the genera *Betacoronavirus* (e.g., SARS-CoV-2, HKU1) as well as human metapneumovirus, human parainfluenzavirus (*Respirovirus*) were detected more frequently and with higher genome completeness in indoor air compared to the wastewater (Figure 2A). In contrast, wastewater yielded more frequent detections and generally higher genome completeness for enteric viruses, particularly members of the *Kobuvirus*, *Salivirus*, and *Sapovirus* virus families (Figure 2B). Notably, some other enteric viruses such as rotavirus A, astrovirus *(Mamastrovirus)* and norovirus were repeatedly detected in indoor air, and several detections reached over seventy-five percent of genome completeness.

**Figure 2.**
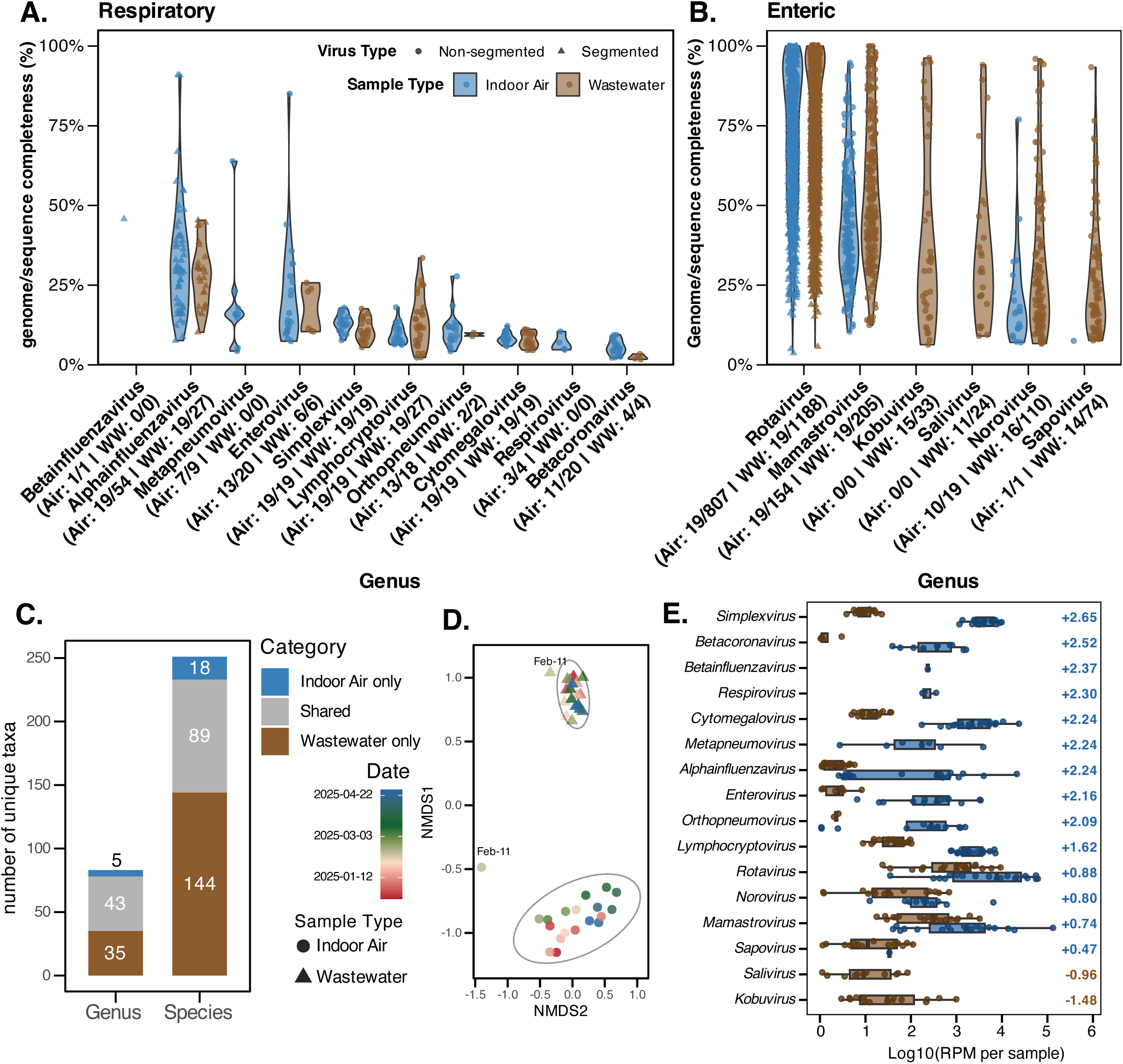
Virus composition and diversity across matched indoor air and wastewater samples (n = 19 matched weeks; 38 total samples). (A–B) Genome completeness (%) distributions by genus for respiratory (A) and enteric (B) viruses; violins are shown per surveillance sample type, and points represent individual detections (genome or gene segments) passing quality filters. For each genus, the number of samples and the number of virus genomes generated for those genera was written in the following parentheses (e.g., *Alphainfluenzavirus* (Air: 19/54) means in 19 samples, 54 segments/sequences were identified). **(C)** Exclusive vs. shared taxa by sample type (unique genera/species across the matched dataset). **(D)** Non-metric multidimensional scaling (NMDS) ordination of species-level composition (Bray–Curtis on within-sample relative abundance; 2D NMDS; stress = 0.084); each point represents one weekly sample (n = 19 per sample type) and ellipses indicate group dispersion. Differences in multivariate dispersion between sample types were tested using PERMDISP (*betadisper* with permutation test; 999 permutations): F(1,36) = 32.99, P = 0.001; mean distance to centroid was 0.40 for indoor air and 0.23 for wastewater. **(E)** Genus-level abundance comparison (log10 summed RPM per sample per genus) between sample types; genera are ordered by median log fold change (median log10[Air] − median log10[Wastewater]). This analysis only compares abundances when both samples have detections.

In terms of exclusivity of certain taxa to sample types, 144 species/35 genera were unique to wastewater, including the known human pathogen parvovirus B19 and numerous animal-associated taxa (e.g., *Aveparvovirus, Iotapapillomavirus*) that are not linked to human infection (Supplementary Figure 1). Conversely, 89/43 taxa were identified across both sample types, while 18/5 were strictly exclusive to indoor air (Figure 2C, Supplementary Figure 1).

To analyze sample-to-sample variability, we used Bray-Curtis distance on species relative abundances per sample. Non-metric multidimensional scaling (NMDS) ordination indicated greater variability in indoor air, with higher dispersion than wastewater (mean distance 0.40 compared with 0.23; betadisper p=0.001; Figure 2D), consistent with more episodic viral community changes in indoor air. Genus-level relative abundance (reads per million filtered reads) comparison of two sample types broadly supported the genome completeness patterns (Figure 2A-B), showing that respiratory viruses yielded more reads per million filtered reads (RPM) in indoor air and most enteric genera (when detected in both) showing comparable RPM across sample types (Figure 2E). However, it should be noted that RPM can be highly influenced by the diversity present in a sample. Less DNA/RNA background (e.g., non-targeted viruses, bacteria) content in indoor air may result in better enrichment for a lower (but targeted) number of viruses, followed by increased post-capture genome amplification and subsequent higher sequencing depth for the fewer viruses found in indoor air.

### 3. Local clinical influenza A cases can be traced with both sample types

Paired influenza A qPCR and metagenomics measurements in indoor air and wastewater enabled a direct comparison of targeted and metagenomics-based surveillance against weekly hospital influenza A positives at University Hospitals Leuven (Figure 3A). Using individual weekly clinical observations, the metagenomics relative abundance signal showed limited agreement with clinical counts, with indoor air not reaching significance (r=0.44, p=0.069) and wastewater showing a modest correlation (r=0.48, p=0.045) (Supplementary Figure 2). Across both sample types, targeted qPCR provided more consistent agreement with clinical positives than metagenomic abundance (indoor air r=0.66, p=0.0027; wastewater r=0.67, p=0.0024; Supplementary Figure 2). Because metagenomics abundance is relative and inter-week variability is substantial (particularly in indoor air metagenomics), we additionally evaluated 4-week rolling averages as a sensitivity analysis for metagenomics and qPCR for both sample types. As expected, rolling averages reduce inter-week stochasticity in relative metagenomic abundance and improved correlation with clinical data, with wastewater metagenomics showing the strongest association (Figure 3B to E).

**Figure 3.**
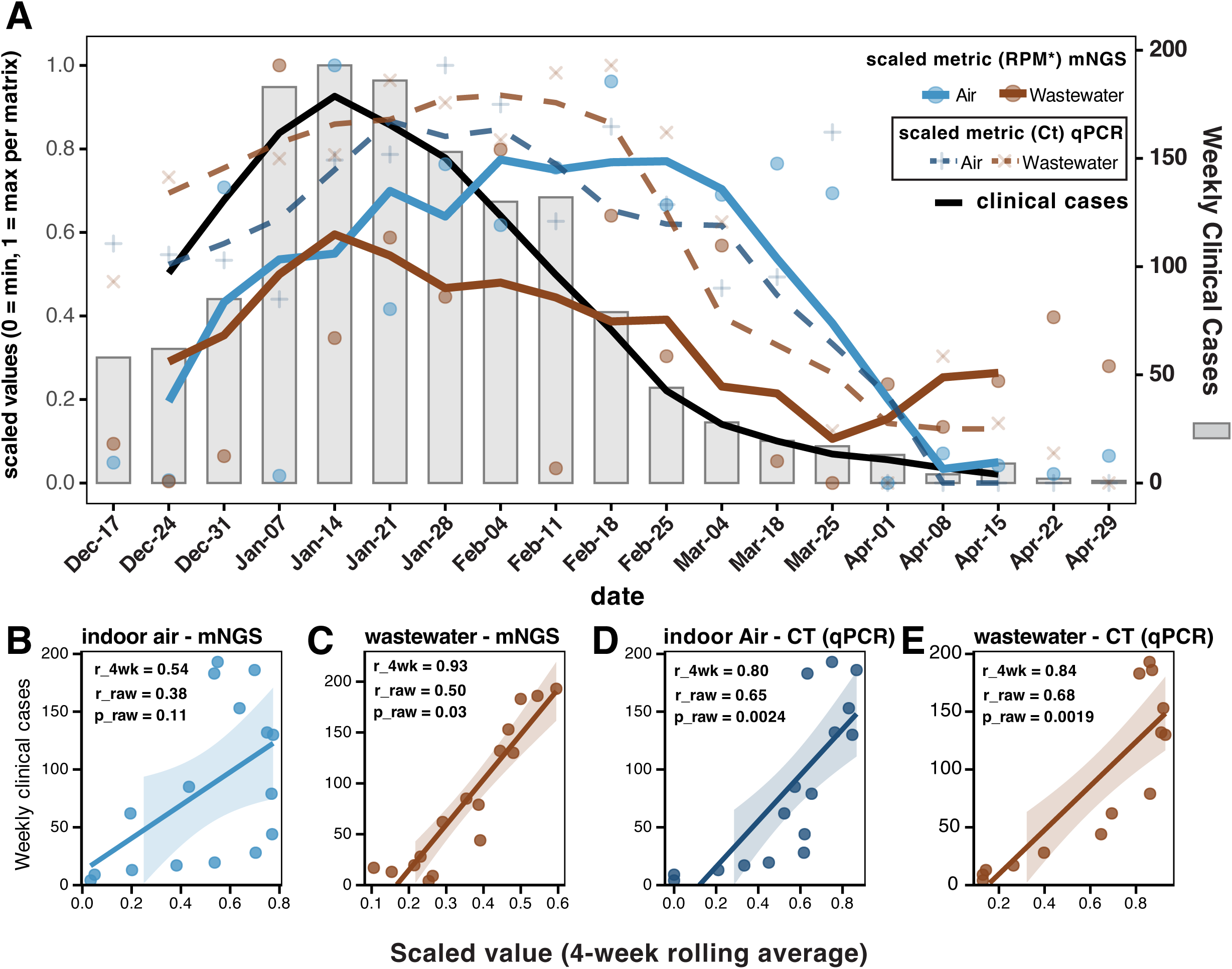
Influenza A dynamics across clinical data, indoor air, and wastewater. **A.** Time series of scaled influenza A abundance from metagenomics in indoor air and wastewater. Spheres show weekly RPM (reads per million filtered reads), and solid lines show their 4-week rolling averages. Indoor air RPM was log10 transformed prior to scaling from 0 to 1, while wastewater RPM was scaled without log10 transformation. Scaled qPCR Ct values are shown as crosses and the 4-week rolling average as dashed lines, where Ct=40 maps to 0 and the lowest observed Ct maps to 1 (indoor air minimum 32.5; wastewater minimum 33.8). Weekly hospital influenza A positives at University Hospitals Leuven are shown as gray bars with a black line (4-week rolling average) on the right axis. **B to E.** Associations between clinical positives and each environmental assay are summarized with scatterplots (x-axis: 4--week rolling average of the scaled environmental signal; y-axis: weekly clinical positives) and linear fits with 95% confidence intervals. Panel annotations report Pearson r for rolling averages (r_4wk), and Pearson r and two-sided p-value computed from unsmoothed weekly values (r_raw, p_raw). Supplementary Figure 2 reports the unsmoothed weekly correlations.

Interestingly, indoor air metagenomics showed an early surge in influenza A abundance, a 2-log increase in reads per million (RPM) from December 24 to December 31 (Figure 3A, blue dot). This early signal occurred one week before clinical cases tripled, yet it was absent in the corresponding qPCR results for those two samples.

### 4. Influenza A subtypes H1N1 and H3N2 cocirculated during the 2024-2025 season and indoor air allowed further in-depth analyses of antiviral resistance mutations in influenza A segments as well as RSV-A fusion protein

Influenza A shows substantial genetic diversity across circulating seasonal lineages, and subtype level resolution is important for interpreting surveillance signals and assessing public health relevance. We therefore performed subtype stratification using competitive mapping against a subtype annotated influenza A reference database. Across the study period, we detected A(H1N1)pdm09 and A(H3N2) in both indoor air and wastewater (Figure 4A to D). A subset of reads remained unassigned, and no other subtypes were detected, consistent with a national surveillance report that predominantly reported A(H1N1)pdm09 and A(H3N2) during this period.^16^

**Figure 4.**
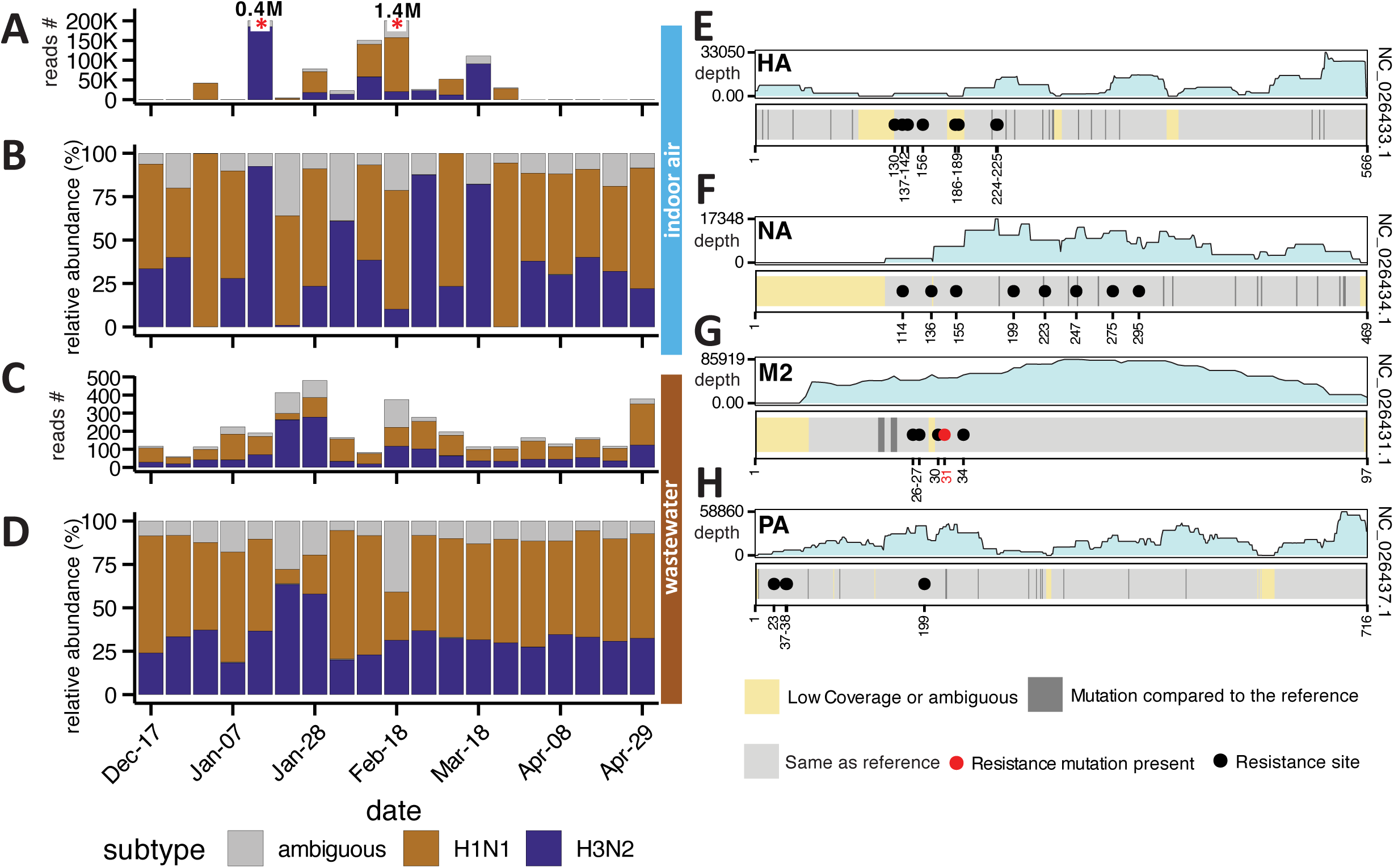
**A-D.** Temporal dynamics of H1N1 (orange) and H3N2 (purple) influenza A subtypes detected in indoor air (A-B) and wastewater (C-D). Read counts (A, C) show total sequence abundance per sample, while relative abundance (B, D) depicts the proportion of each subtype, with grey indicating ambiguous or unclassified reads. *Some samples exceed 200K reads (marked with breaklines and read counts labeled above).* **E-H.** Sequencing coverage and amino acid conservation across four key genes (HA, NA, M2, PA) in a representative H1N1 sample. The upper panels show sequencing depth across each ORF, while the lower panels display the conservation status of each amino acid position. Positions are colored by status: grey indicates identity with the reference genome, light yellow indicates low coverage (< 100 reads) or ambiguous nucleotides, and dark grey indicates mutations relative to the reference. Black dots mark known (checked against) antiviral resistance sites; red dots indicate positions where known resistance-conferring mutations are present in the sample. Vertical lines in each plot at the bottom of every panel indicate individual amino acid positions. Gene references are indicated on the right (GenBank - RefSeq accessions).

We next verified whether we could screen for antiviral resistance markers in influenza A segments when coverage was sufficient. Wastewater influenza A genome recovery remained too fragmented (<50% coverage) for mutation assessment. In contrast, indoor air yielded sufficient coverage in the highest-coverage A(H1N1)pdm09 indoor air sample (February 18, Supplementary Table 1 for focused resistance sites). While we did not identify the most common mutation H275Y in N1 for Oseltamivir, we identified the well-established M2 substitution S31N (Figure 4E to H), which is associated with adamantane resistance and is carried by more than 99 percent of circulating A(H1N1)pdm09 strains^17^. Beyond S31N, we did not detect meaningful minority variants at resistance associated sites, with all alternative alleles below 1 percent at positions evaluated (Supplementary Table 2). Additional amino acid differences relative to the reference sequence are listed in Supplementary Table 3.

Lineage assignment of influenza A viruses can also guide clinical and public health decisions. Given recent attention to the emergence of H3N2 K lineage viruses in 2025^18^, we examined the HA consensus sequence for the highest coverage A(H3N2) sample, however this did not show evidence for the presence of the K lineage for that particular sample. Clade assignment using Nextclade (version 3.18.1)^19^ placed this sequence within clade J.2, close to J.2.5 (Supplementary figure 3).

In light of the increasing use of monoclonal antibodies for RSV prophylaxis in Belgium, we additionally evaluated respiratory syncytial virus A for clesrovimab resistance markers. We recovered the fusion protein region spanning residues 397 to 551, confirming the absence of known resistance substitutions, although incomplete coverage precluded full-gene resistance assessment (Supplementary Table 4 for resistance mutations analyzed).

### 5. Indoor air surveillance coupled with metagenomics can help wastewater surveillance on decrypting the surveillance patterns of ignored viruses

Many human associated viruses show distinct tissue tropism and shedding routes, which can complicate interpretation of an environmental surveillance signal. This is particularly relevant for wastewater, where signals reflect a composite of human and animal inputs and are shaped by strong sample specific effects on relative abundance. We therefore used *Mastadenovirus* and *Enterovirus* as two examples to provide evidence of how paired indoor air and wastewater metagenomics can show sample type specific patterns (Figure 5A to H).

**Figure 5:**
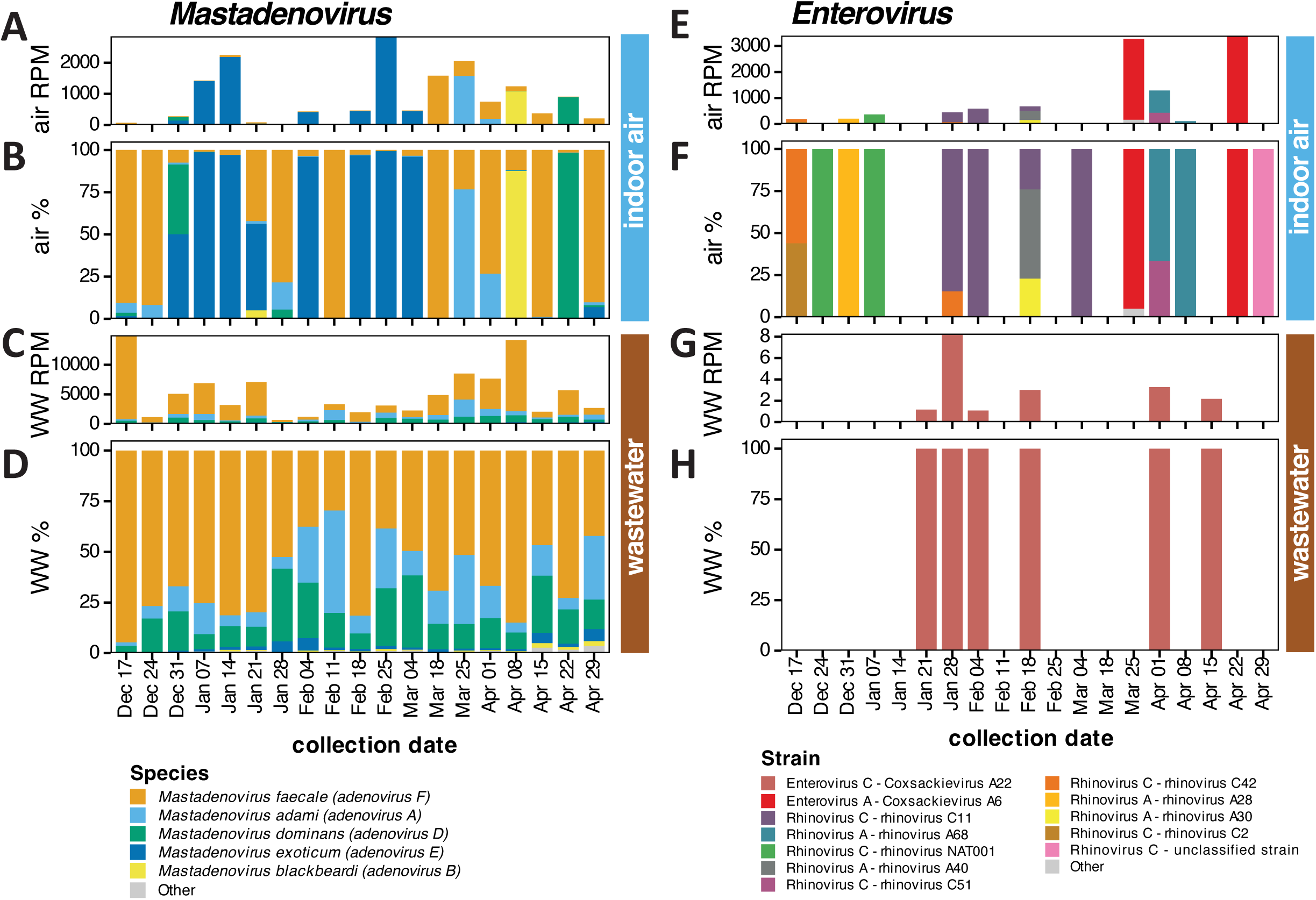
Relative abundance of viral species per genus in paired indoor air and wastewater samples. Stacked bar plots showing the composition of adenoviruses (A–D) and enteroviruses (E–H) detected across matched pairs of indoor air and wastewater samples collected in the same week. (A, E) Viral abundance in indoor air (reads per million: RPM). (B, F) Relative species or strain composition in indoor air (%). (C, G) Viral abundance in wastewater (RPM). (D, H) Relative species or strain composition in wastewater (%).

Adenoviruses comprise over 50 serologically distinct types across seven human species (A-G) and cause diverse clinical manifestations, from respiratory infections to gastroenteritis (mostly but not limited to species F) depending on the serotype.^20^ Indoor air was dominated by human adenovirus 4 (species mastadenovirus E), a pathogen almost exclusively associated with respiratory infections and outbreaks. ^20^ In contrast, this respiratory species contributed <5% to the wastewater adenovirus community, which was instead dominated by species F, a group classically associated with gastroenteritis (Figure 5A to D).

Enteroviruses consist of 250 serotypes causing clinical presentations ranging from mild respiratory illness (most rhinoviruses) to severe neurological complications (e.g., poliovirus).^21^ The respiratory Rhinovirus A and C, alongside Coxsackievirus A6 (a known cause of hand, foot, and mouth disease), were detected exclusively in indoor air.^22,23^. Conversely, wastewater consistently captured Coxsackievirus A22, a subtype rarely identified in routine clinical surveillance. Moreover, rotavirus species, norovirus subtypes, multiple papillomaviruses as well as astrovirus strains could be followed up in both indoor air and wastewater, showing predominantly matching genetic imprint in both sample types (Supplementary Figures 4-5).

## Discussion

Wastewater studies typically report broad viral diversity and stable temporal patterns ^5,24^, whereas indoor air studies mostly emphasize location-specific, episodic respiratory detection.^6–8^ Because the two sample types represent different surveillance units and integration windows, our study design primarily enables a within-season comparison of how signals differ across sample types under a common wet-lab processing and analyses workflow, rather than a direct head-to-head estimate of community incidence. Overall, we show that wastewater is valuable for One-Health approaches and population monitoring, while indoor air provides a sentinel, human-centered and respiratory virus view in a targeted setting.

Consistent with its nature as an integrative, composite sample gathering animal and human waste^6,7,10^, wastewater yielded substantially higher viral diversity and more animal-associated viruses than indoor air. Conversely, indoor air provided a more “human-oriented” respiratory virus focus. This is particularly relevant to highly pathogenic avian influenza (HPAI) which is not detected in our current study, while it has been reported in wastewater across multiple regions yet cannot be directly linked to human circulation.^8,9,27,28^ Indoor air surveillance in sentinel clinical settings could complement wastewater signals by adding a human-respiratory context, helping distinguish likely human circulation from non-human or environmental contributions when wastewater detections are ambiguous.

As demonstrated by the high sample-to-sample variability in our data, viral signals in indoor air are episodic, likely reflecting acute events driven by individual differences in viral shedding in an emergency department microenvironment, a dynamic supported by existing indoor air studies.^11–13,25^ Notably, our probe-based hybridization approach achieved >75% genome completeness for several respiratory and enteric viruses in indoor air in multiple instances (i.e., rotavirus, astrovirus, and norovirus). This observation motivates follow-up work to understand the mechanisms contributing to the detection of enteric viruses in indoor air, such as toilet-derived aerosolization^29^ and/or possibly saliva-associated shedding.^30^ It is crucial to note that our data cannot distinguish the relative contribution of these routes and infectivity of the viruses identified.

Our high rates of genome completeness highlight the value of hybrid-capture viral metagenomics for both low-biomass (indoor air) and high-background (wastewater) sample types. While earlier environmental studies using untargeted shotgun sequencing often reported fragmented viral genomes, our targeted enrichment resulted in high genome completeness for many viruses across both sample types, consistent with findings in other complex sample types.^12,13,31^. However, outcomes remain sample-dependent: the clinical indoor air environment provided higher human-specificity. Therefore, we suggest prioritizing indoor air when the goal is to minimize background complexity and maximize sequence quality for human-associated respiratory viruses.

Existing literature suggests that indoor sampling can track epidemiology of viruses^26,32^ and a previous study identified coronavirus HKU1, human parainfluenza virus 3 and 4 outbreaks concurrently or a week in advance to the increase in clinical cases.^26^ We confirm possibility of tracking epidemiology in principle; however, the one-week early signal of influenza A we observed via indoor air metagenomics was absent when targeted qPCR was used, suggesting it may reflect the high sample-to-sample variability inherent to the sequencing approaches. Nonetheless, hybrid-capture enabled influenza A subtyping and characterization in both sample types. When coverage thresholds were met, we successfully conducted targeted resistance-site screening for both influenza A and RSV-A from indoor air, suggesting possibility of vaccine-preventable pathogen monitoring in high-risk populations.

All in all, resistance monitoring with indoor air metagenomics may not be feasible in every sample due to variable genomic coverage, a limitation that must be factored into the design of future environmental surveillance studies. In our study, raw samples were stored at -80°C up to 5 months before nucleic acid extraction for both sample types. Literature suggests that enveloped respiratory viruses, particularly influenza A and RSV, are more susceptible to freeze-thaw degradation compared to non-enveloped enteric viruses, with RSV detection rates dropping dramatically after freezing. ^33^ This differential stability may explain the incomplete genome recovery observed for influenza viruses and RSV in indoor air samples, even when read counts were high. To optimize recovery for resistance monitoring, we recommend immediate nucleic acid extraction after collection, to minimize freeze-thaw degradation.

Overall, we propose a layered environmental surveillance strategy utilizing two complementary lenses: wastewater provides a stable, highly diverse backbone for population-level One Health sensitive, wide-angle lens; while indoor air adds a targeted, focused lens for human environments. In zoonotic scenarios such as HPAI, wastewater can flag these signals early on a population level, whereas indoor air metagenomics at clinical interfaces provides critical confirmation of human circulation and enable genomic characterization when genome coverage permits. This dual approach is especially relevant for global surveillance, as wastewater signals in low- and middle-income countries or regions with open sanitation are heavily influenced by animal waste and agricultural runoff, making the human-specific confirmation critical for detecting true zoonotic spillover.

Our study has several limitations warranting careful interpretation. First, it was conducted in a single city during one influenza season, which constrains generalizability across seasons, locations, and building types. Second, indoor air and city-wide wastewater represent fundamentally different surveillance units, with indoor air reflecting a week-long, location-specific sample while wastewater representing a 24-hour composite of a city-scale population. Third, detection of viral nucleic acids in wastewater and indoor air does not imply viral viability, infectiousness, or transmission risk, nor does it allow attribution to specific aerosolization or exposure routes for indoor air. Fourth, the use of hybrid-capture metagenomics introduces reference-dependent biases that can influence apparent diversity and relative abundance even though probes can bind up to 20% divergence reportedly. Despite these limitations, our findings demonstrate that pairing the broad, population-level scope of wastewater monitoring with the targeted, human-centric specificity of indoor air sampling provides a highly complementary approach for future genomic surveillance approaches.

## Methods

### Wastewater sampling and processing

Wastewater sampling^34^ and processing followed previously established protocols used for longitudinal viral metagenomic surveillance^35^ in Leuven. Briefly, 24-hour composite influent samples were collected at the municipal wastewater treatment plant using time-proportional sampler, sampler collecting 50 mL every 10 minutes. Samples were transported at 4 °C and stored at -80C until DNA/RNA extraction and probe-based hybridization assisted viral metagenomics, which was conducted in May 2025 (1 month to 5 months after sample collections in this study).

### Indoor air sampling via HVAC system and qPCR testing

Indoor air sampling was adapted from a centralized HVAC return-air sampling strategy previously described ^32^ for respiratory virus surveillance, with modifications to support continuous, week-long sampling in a hospital emergency department. Sampling was conducted weekly between December 12, 2024 and April 29, 2025. Following collection, air sampling cartridges were sealed, transported at room temperature and processed using the multiplex qPCR panel Alinity m resp-4-plex assay (Abbott, Illinois, USA), targeting SARS-CoV-2, influenza A/B and RSV. Negative controls were included to monitor contamination during processing in the lab. Left-over samples after qPCR were stored at -80C until processing with probe-based hybridization assisted viral metagenomics together with wastewater samples. Air was sampled from the HVAC exhaust plenum (see supplementary methods for the placement of air sampler, airflow rates and sample processing), where extracted air from all rooms of the emergency department was mixed. Samples were collected using a high-flow aerosol sampler (Thermo Fisher Scientific, AerosolSense), as explained before. ^32^

### Library preparation, hybrid capture, and sequencing statistics

To ensure comparability between sample types, identical library preparation, hybrid-capture enrichment, and sequencing workflows were applied to indoor air and wastewater samples, as explained before.^35^ Libraries were prepared from extracted nucleic acids, pooled up within sample types (i.e., indoor air samples and wastewater samples pooled separately, 19 samples and a negative control for each group) up to 4000 ng of nucleic acid material and enriched using the Comprehensive Viral Research Panel (TWIST Biosciences, USA) reportedly targeting more than 3,000 human and animal infecting virus species. Enriched libraries were amplified for 13 cycles and sequenced on the AVITI platform (Element Biosciences, USA) using 150 bp paired-end reads.

Sequencing yielded a mean of 17.0 million quality-filtered reads per indoor air sample (SD: 3.9M) and 31.1 million reads per wastewater sample (SD: 7.6M). Our analyses focusing on rarefaction and species accumulation demonstrated that taxon discovery reached saturation well below our sequencing depth per sample. Full details of these ecological validation steps and statistical models are provided in the Supplementary Methods.

### Bioinformatics analyses

Metagenomics analyses were conducted using EsViritu (v.0.2.3)^5^. Briefly, sequencing reads were quality filtered, and adapter trimmed, and deduplicated prior to viral analysis. Viral identification was performed using reference-based mapping against a curated viral genome database (Virus Pathogen Database for EsViritu, record 7876309 on Zenodo). After detection and calculation of abundance and covered bases, additional filters were applied to keep only true positive consensus sequences: reads per million (RPM) filtered reads >1, covered bases >500 bp, while for genomes larger than 100 kb (e.g., *Herpesviridae*) at least 3000 bp needed to be covered. Detected viral genera were categorized into two primary functional groups (respiratory and enteric viruses) based on primary transmission routes and clinical relevance. A complete breakdown of these classifications is provided in the Supplementary Methods.

### Influenza subtype and antiviral resistance analyses

Influenza A subtypes were determined by competitive mapping of reads to a curated subtype annotated- reference database (influenza A serotype workflow as described previously)^9^. Reads that did not support a unique subtype assignment were retained as “ambiguous” to avoid forcing classifications.

For antiviral resistance surveillance, resistance-associated amino acid positions were evaluated only in genomic regions meeting coverage depth requirements (>100 reads), while resistance mutations outside informative genomic regions cannot be excluded. The full reference set, inclusion thresholds, and curated resistance site lists are provided in the Supplementary table 1 and scripts.

### Quantitative PCR and clinical data integration

Quantitative PCR (qPCR) data for influenza A virus were used as an orthogonal comparator to metagenomic signals. Ct values were scaled to a 0–1 range within each sample type to facilitate comparison with abundance measures. For wastewater, direct qPCR data was unavailable for our sequenced samples; therefore, we retrieved qPCR data from independent wastewater samples^36^ collected during the corresponding weeks from the publicly available Leuven wastewater surveillance dashboard for comparison.^37^ For indoor air, qPCR was directly conducted from the same sample as explained above and used for comparison. Clinical case numbers for comparisons were derived from the publicly available dashboard of University Hospitals Leuven for respiratory infections, which includes positive tests from the admitted patients, outpatient tests and external tests.^15^

### Statistical analyses

Viral community composition was compared across sample types using genome completeness and within-sample relative abundance, restricting anlyses to quality-filtered detections and matched weekly sample pairs. Beta diversity was quantified using Bray–Curtis dissimilarity on species-level relative abundance tables and visualized by NMDS; differences in multivariate dispersion between sample types were tested using PERMDISP (betadisper with permutation test; details in the Supplementary Methods).

Associations between environmental influenza A signals and weekly hospital influenza A positives were summarized using two-sided Pearson correlations. Metagenomic abundance and qPCR Ct were scaled within each sample type for comparability (with indoor-air metagenomic RPM log10-transformed prior to scaling). For inference, *p*-values were computed from unsmoothed weekly values and *p*-value below 0.05 was deemed significant. Statistical analyses were conducted in R (version 2023.06.0+421); scripts and full settings are provided in the code repository.

## Data Availability

Sequencing data have been deposited in SRA (Sequencing Read Archive) under accession number PRJNA1431177. Custom scripts used for data processing and analysis are available at https://github.com/Matthijnssenslab/air_wastewater_leuven/.

## Ethical statement

The study was approved by the Ethics Committee Research UZ/KU Leuven (S66518). No informed consent was required from occupants of the sampled environments.

## Acknowledgements

Mustafa Karatas is supported by a Research Foundation Flanders (FWO) fundamental research scholarship (number: 11P7I24N). C.G., L.C., E.H., S.G. and E.A. acknowledge support from the DURABLE project. The DURABLE project has been co-funded by the European Union, under the EU4Health Programme (EU4H), project no. 101102733. Views and opinions expressed are however those of the authors only and do not necessarily reflect those of the European Union or the European Health and Digital Executive Agency. Neither the European Union nor the granting authority can be held responsible for them. The computing power in this work was provided by the VSC (Flemish Supercomputer Centre), financed by the FWO and the Flemish government–department EWI.

## Use of artificial intelligence tools

ChatGPT-5.2 and Gemini 3.1 Pro have been used for text and Rscript proofreading purposes.

