## Supplementary methods for "A Tale of Two Lenses: Emergency department indoor-air hybrid-capture metagenomics complements wastewater by adding a human-focused respiratory virus perspective"

**Wastewater sample concentration and extraction**

Influent wastewater samples consisted of 24-hour time-proportional composite influent collected at the Leuven municipal wastewater treatment plant, transported at 4 °C, and stored at −80 °C until laboratory processing (processed together with the air samples in May 2025). For each sample, 450 mL of influent wastewater was concentrated by stirred cell ultrafiltration (UFSC40001, Merck Amicon) using a 30 kDa ultrafiltration membrane (Ultracel 30 kDa; PLTK07610, Merck Millipore) under compressed air. The retained fraction was recovered in 4.5 mL nuclease-free water (100× concentrate). The concentrate was clarified by centrifugation at 10,000 × g for 10 min, and the supernatant was used for downstream analyses. Nucleic acids were extracted from 350 µL of the wastewater concentrate using the RNeasy Mini Kit (Qiagen; 74106), following the manufacturer’s instructions. A negative extraction control (350 µL sterile water) was processed in parallel with each extraction batch. Extracted nucleic acids were eluted in 50 µL elution buffer and stored at −80 °C until downstream library preparation and sequencing (described in the main Methods). Because targeted qPCR was not performed directly on the aliquots used for metagenomic sequencing, we utilized publicly available longitudinal qPCR data from the Leuven wastewater surveillance program as a comparative proxy. To ensure temporal relevance, the external qPCR data paired with our metagenomic results were restricted to independent samples collected within the exact same epidemiological week as our sequenced aliquots.

Drainage area of the wastewater treatment plant can be found in our previous study: “Karatas, Mustafa and Bloemen, Mandy and Swinnen, Jill and Matthijnssens, Jelle and PDF, See, Hybrid-Capture-Enabled Longitudinal Metagenomics Allows Strain-Resolved Human-Associated Virus Surveillance in Wastewater. Available at SSRN: <https://ssrn.com/abstract=6047769> or [http://dx.doi.org/10.2139/ssrn.6047769](https://dx.doi.org/10.2139/ssrn.6047769)”.

**Indoor air sampling via HVAC system**

Indoor air sampling was performed using a centralized sampling strategy targeting the heating, ventilation, and air conditioning (HVAC) air, as described previously (Happaerts et al., 2024; Raymenants et al., 2023). This approach allows for the scalable surveillance of a large, multi-room environment by sampling at the confluence of extracted air.

A high-flow active air sampler (AerosolSense™, Thermo Fisher Scientific) was installed in the HVAC return plenum of the University Hospitals Leuven emergency department (**Figure 1, below**). This sampling location represents a mixing point where extracted air from all patient waiting areas, clinical zones, and restrooms within the department converges. Sampling was conducted continuously approximately for one week per sample between December 12, 2024, and April 29, 2025. The sampler operated with the standard AerosolSense cartridges at a fixed flow rate of 200 L/min.

Following collection, the sampling cartridges were removed, sealed, and transported to the laboratory at room temperature. To elute the captured aerosols, the two collection substrates were removed from the cartridge and incubated together in 4mL of viral transport medium, and  vortexed for 30 seconds before removing the substrates. A 500 µL aliquot of this eluate was used for immediate initial screening using the Alinity m resp-4-plex assay (Abbott, Illinois, USA) to detect SARS-CoV-2, influenza A/B, and RSV. The remaining eluate was stored at -80°C until nucleic acid extraction and library preparation for viral metagenomics, which was performed in parallel with wastewater samples.

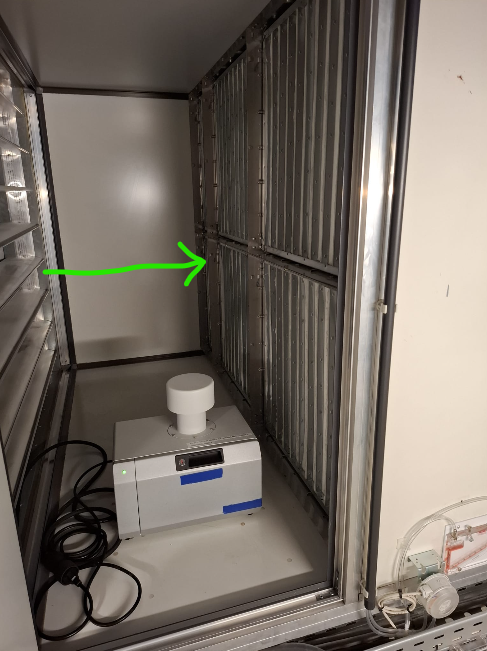

Figure A. Thermofisher AerosolSense sampler in the HVAC plenum. Green arrow shows air flow from emergency department rooms to the central filtering system, downstream from the sampler.

**Bioinformatics analyses**

**Data filtering and quality control**

To ensure high-confidence viral detections, we applied a multi-step filtering approach (script: *filter_data.R* in the GitHub repository). Detections were retained only if they met the following criteria: (1) a minimum of 10 aligned reads, (2) an abundance of >1 read per million (RPM), and (3) either a genome coverage $\geq$ 500 bp or $\geq$ 50% detected genome completeness. Known laboratory contaminants (e.g., *Parvovirus NIH-CQV*, *Chikungunya virus*) and spurious hits (e.g., *Alphamesonivirus*, *Retroviridae*) were explicitly removed. Additionally, we filtered out short amplicon-like signals by requiring *Human mastadenovirus C* detections to have $\geq$ 20% genome coverage (These viruses were identified in the negative control samples in the current run and previous sequencing runs).

**Respiratory and enteric virus classification**

To contextualize the viral detections within an aerobiological and wastewater surveillance framework, we categorized detected viral genera into two functional groups based on their primary transmission route and clinical relevance:

1. **Respiratory Virus Group:** This category includes viruses primarily known for respiratory transmission (aerosols/droplets) or those commonly detected in respiratory tract infections. This group encompasses:
   - **“Classic” respiratory pathogens:** *Orthopneumovirus* (RSV), *Metapneumovirus*, *Alphainfluenzavirus* (Influenza A), *Betainfluenzavirus* (Influenza B), *Respirovirus* (Parainfluenza), and *Betacoronavirus*.
   - **Respiratory/Oral-Shed Viruses:** *Enterovirus* species were classified in this group due to the inclusion of rhinoviruses and respiratory enteroviruses (e.g., EV-D68) which are highly relevant for air sampling.
   - **Saliva/Oral-shed and pathogens with respiratory transmission potential:** Several *Herpesviridae* genera (*Cytomegalovirus*, *Lymphocryptovirus* [EBV], *Simplexvirus*) were also retained in this category as they are shed in oral secretions and can be aerosolized, despite that their infection symptoms may not always be respiratory.
2. **Enteric Virus Group:** This category includes viruses classically associated with the fecal-oral transmission route and gastrointestinal infection, which are the primary targets of standard wastewater surveillance. This group encompasses: *Norovirus*, *Rotavirus*, *Mamastrovirus* (Astrovirus), *Sapovirus*, *Kobuvirus*, and *Salivirus*. We acknowledge that some respiratory viruses may multiply in enteric tract therefore this classification may not be definitive, and vice-versa may be correct for enteric viruses.

**Sequencing related statistics**

Indoor air samples (n = 19) yielded a total of 322,794,394 filtered reads (after QC as explained in the methods section), with a mean of 16,989,179 filtered reads per sample (median: 16,850,176; SD: 3,877,112; range: 12,661,044–27,023,440). Wastewater samples (n = 19) yielded a total of 590,670,410 filtered reads, with a mean of 31,087,916 filtered reads per sample (median: 30,468,607; SD: 7,625,893; range: 15,336,139–50,204,723). On average, indoor air samples yielded 51.8 viral species per sample (median: 52), compared to 113.9 species per sample (median: 115) in wastewater.

**Sequencing depth, rarefaction, and species accumulation analyses**

To assess sequencing depth sufficiency and compare viral diversity capture between indoor air and wastewater, we performed ecological diversity analyses using R with the *vegan* package. To estimate the total expected viral richness in each environment, we generated species accumulation curves using the specaccum function (method = "random" with 999 permutations). This method plots the cumulative number of unique viral species discovered as a function of the number of samples analyzed, averaged over random orderings of the samples. The curve suggests processing samples may potentially result in identification of more species (Figure S2A).

Next, we evaluated the relationship between sequencing effort (total filtered reads) and observed alpha diversity (number of viral species detected) for each sample (Figure S2B), since this can be relevant to the analyses we conducted. Pearson and Spearman correlations were calculated to test the strength of the association. Additionally, linear regression models were fitted separately for indoor air and wastewater samples to determine if higher sequencing depth significantly predicted increased species recovery, which shows that higher sequencing depth did not result in higher number of species in our sample types ($p>0.05$). This is possibly due to the nature of hybrid-capture’s selection process as well as our sample matrix only containing limited number of viruses, which, possibly we have sequenced it all.

To determine if the sequencing depth was sufficient to capture the majority of the viral diversity (species and strains), we performed rarefaction analysis on the pooled reads from all samples within each sample type using *rarecurve* (Figure 2C-D). Reads were subsampled at intervals of 1/1,000th of the minimum total library size. We defined the **saturation point** as the sequencing depth where the accumulation of newly discovered taxa dropped below 1 new taxon per 1 million additional reads (slope $\left< {10}^{-6} \right.$). As shown in dashed lines, after sequencing for 0.6M (for species) and 1.6M (for strains) in indoor air detections saturated. For wastewater, saturation points were 1.9M for species detection and strain detection was at 2.4M filtered total reads per sample.

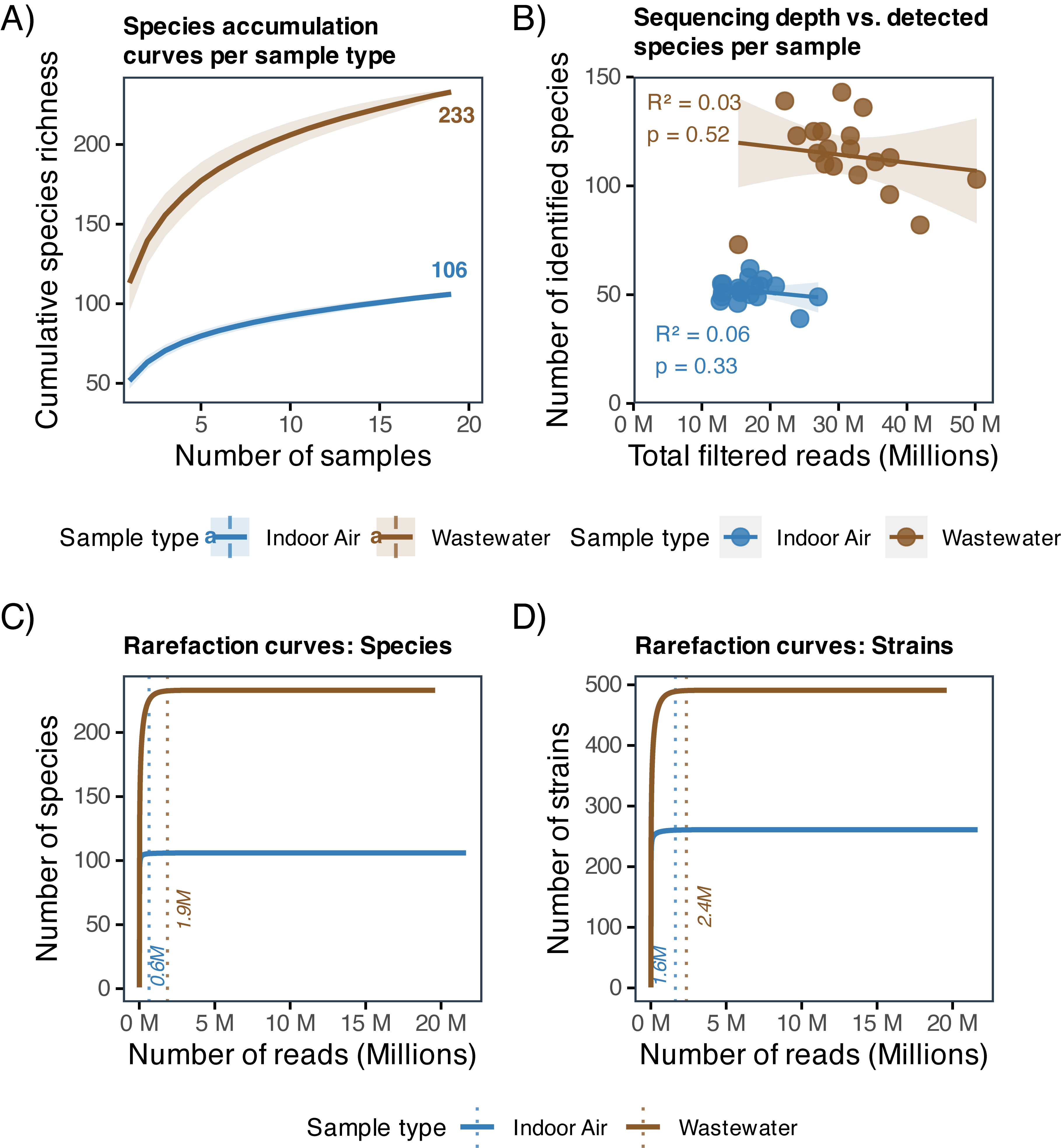

**Figure S2. Evaluation of sequencing depth for viral detection in indoor air and wastewater.**

**(A)** Species accumulation curves displaying the cumulative number of unique viral species detected as sample size increases. Curves represent the mean richness over 999 random permutations of sample order; shaded bands indicate standard deviation. The final labels show the total species richness observed in the full dataset for each environment. **(B)** Scatter plot of sequencing depth (in millions of filtered reads) versus the number of detected viral species per sample. Lines represent linear regression fits with 95% confidence intervals (shaded). Goodness-of-fit ($R^{2}$) and $p$-values are provided for each sample type in the figure. **(C–D)** Rarefaction curves for viral **(C)** species and **(D)** strains based on pooled reads from all samples per environment. The curves show the expected number of distinct taxa detected at varying subsampling depths. Dotted vertical lines indicate the saturation point, defined as the sequencing depth beyond which fewer than one new taxon is expected per one million additional reads. The values on the dashed lines show the depth (in millions of reads) required to reach this saturation threshold.

**Evaluation of genome completeness and differential abundance metrics**

Genome completeness distributions were compared between sample types to assess the integrity of viral genomes recovered from each matrix. We calculated the percentage of the reference genome covered by mapped reads for every valid detection. Violin plots were generated to visualize the density distribution of genome completeness values for each viral genus. In these visualizations, individual detections (segments or complete genomes) were overlayed as points to show the raw data distribution alongside the kernel density estimation.

To identify viral taxa that were enriched or exclusive to a specific surveillance medium, we calculated the **Log Fold Change (LFC)** of normalized abundance (RPM). For genera detected in the dataset:

1. Abundance counts were RPM-normalized to control for library size differences.
2. Values were $\log_{10}$-transformed to stabilize variance.
3. The median $\log_{10}$ RPM was calculated for indoor air and wastewater groups separately.
4. LFC was derived as $\left( \text{Median}_{\text{Air}} - \text{Median}_{\text{WW}} \right)$.
   - Positive LFC values indicate enrichment in indoor air.
   - Negative LFC values indicate enrichment in wastewater.
   - Genera were ranked by LFC to highlight matrix-specific preferences.

**Ecological dissimilarity (Beta diversity, Figure 2D)**

Beta diversity was quantified using  Bray-Curtis dissimilarity on species-level relative abundance tables. This metric was selected because it accounts for both the presence/absence of species and their relative abundance. Unconstrained ordination was performed using Non-metric Multidimensional Scaling (NMDS) ($k=2$, trymax=200) to visualize compositional differences. To confirm that the observed separation was not driven by heterogeneous variances, we performed a *PERMDISP* test (multivariate homogeneity of group dispersions) with 999 permutations, testing the null hypothesis that the average distance of samples to their group centroid is identical between indoor air and wastewater.
