## Supplementary information for "A Tale of Two Lenses: Emergency department indoor-air hybrid-capture metagenomics complements wastewater by adding a human-focused respiratory virus perspective"

**Supplementary information (Figures and tables)**

**Supplementary Figure 1.** Sample type preference of genera identified in the study. Left and middle figures were done based on individual sequence detections, while the left table is based on how many samples contained mentioned genera.

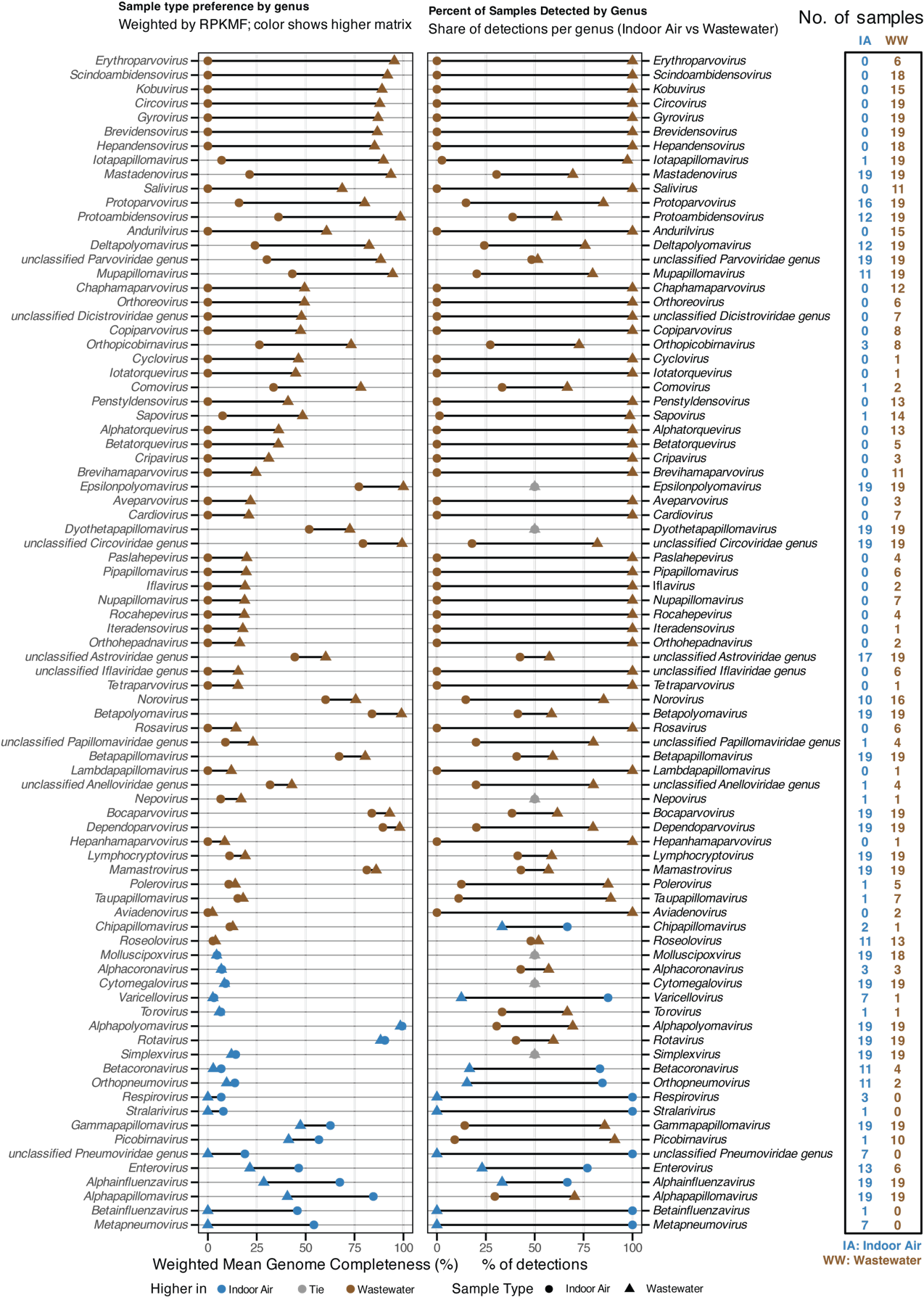

**Supplementary Figure 2.A-D.** Individual weekly values correlation analyses without 4-weekly averages.

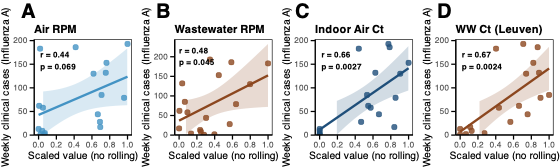

**Supplementary Figure 3.** Clade identification of H3N2 HA gene recovered from one of the samples. Only 42% (309 nt) of HA2 region was used for these analyses using Nextclade online server (version 3.18.1, clades.nextstrain.org) with the dataset: nextstrain/flu/h3n2/ha/EPI1857216 (Influenza A H3N2 HA).

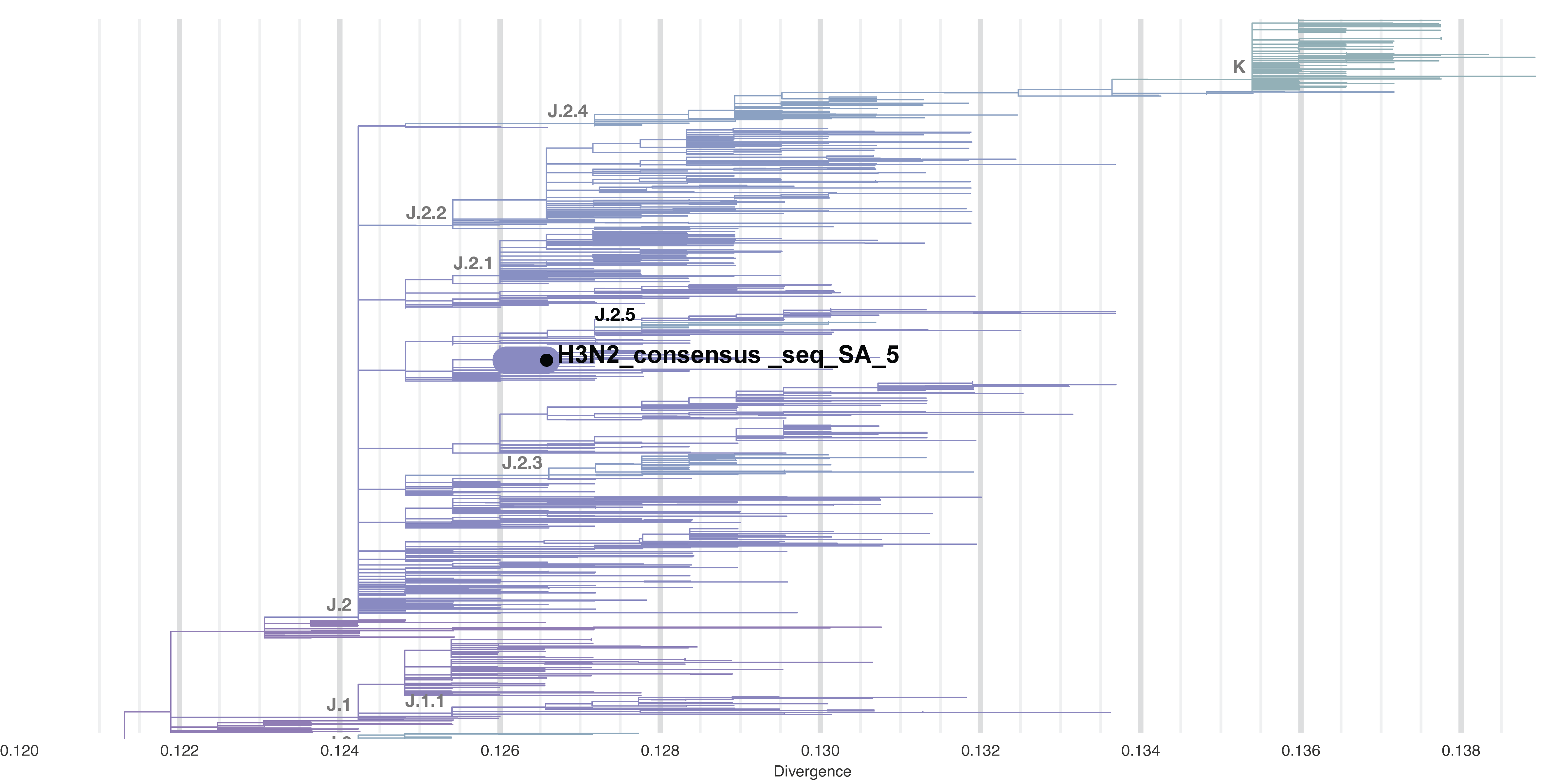

**Supplementary Figure 4.** Relative abundance of additional viral genera in paired indoor air and wastewater samples. Stacked bar plots showing the composition of Mamastrovirus, Norovirus, and Papillomavirus across matched sample pairs. Each color represents a distinct viral strain within each genus (or a genus within a family for the *Papillomaviridae*). Filtering criteria are the same as Figure 5.

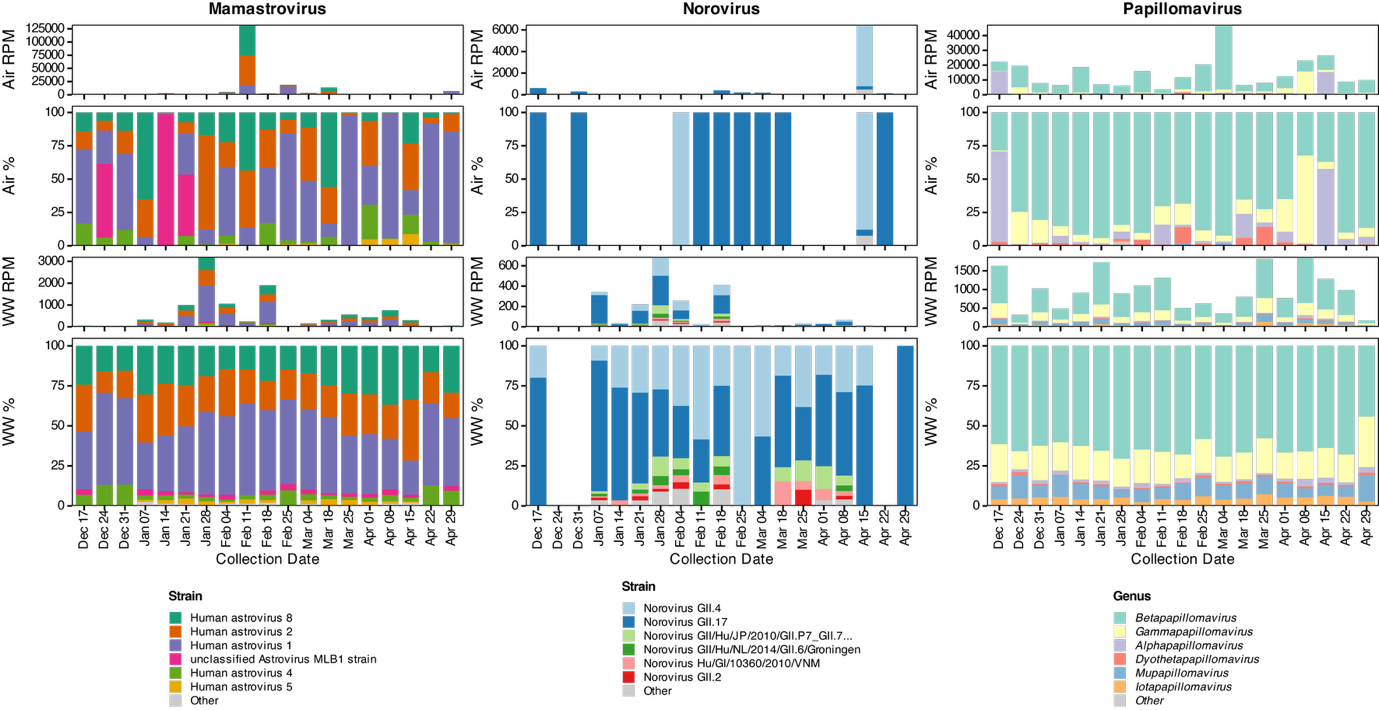

**Supplementary Figure 5. Rotavirus species abundance in paired indoor air and wastewater samples.** Stacked bar plots showing the viral load (reads per million, RPM) of rotavirus species detected in (A) indoor air and (B) wastewater samples collected in the same week.

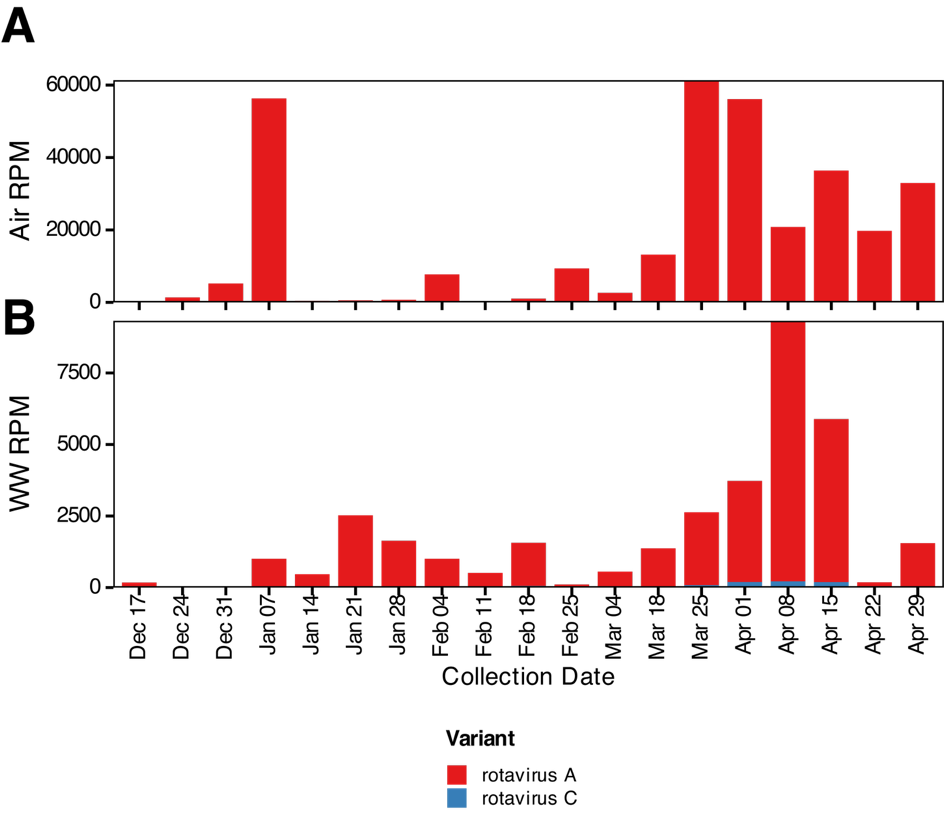

**Supplementary table 1.** H1N1 mutations analyzed in this study and sources

| **Segment** | **Amino acid change** | **Drug target or importance of the mutation** | **Resistance level** | **Source** |
| --- | --- | --- | --- | --- |
| **NA (N1)** | H275Y | Oseltamivir; Peramivir | Highly reduced inhibition (HRI) | <https://www.cdc.gov/flu/treatment/antiviralresistance.html> \| <https://academic.oup.com/jid/article/232/Supplement_3/S169/8287899> |
| **NA (N1)** | I223V | Oseltamivir; Peramivir | Reduced inhibition (RI) | <https://pmc.ncbi.nlm.nih.gov/articles/PMC11600549/> \| <https://pmc.ncbi.nlm.nih.gov/articles/PMC12270646/> |
| **NA (N1)** | I223R | Oseltamivir; Peramivir | Reduced inhibition | <https://pmc.ncbi.nlm.nih.gov/articles/PMC12270646/> |
| **NA (N1)** | I223T | Oseltamivir | Reduced inhibition | <https://pmc.ncbi.nlm.nih.gov/articles/PMC12270646/> |
| **NA (N1)** | S247N | Oseltamivir | Reduced susceptibility | <https://pmc.ncbi.nlm.nih.gov/articles/PMC12270646/> |
| **NA (N1)** | N295S | Oseltamivir | Reduced susceptibility | <https://pmc.ncbi.nlm.nih.gov/articles/PMC11600549/> |
| **NA (N1)** | Q136K | Zanamivir; Peramivir; Laninamivir | Highly resistant | <https://pmc.ncbi.nlm.nih.gov/articles/PMC11600549/> |
| **NA (N1)** | Y155H | Oseltamivir; Zanamivir; Peramivir | 30-100 fold reduced | <https://pmc.ncbi.nlm.nih.gov/articles/PMC3772742/> |
| **NA (N1)** | V114I | Oseltamivir; Zanamivir; Peramivir | Reduced susceptibility (small) | <https://pmc.ncbi.nlm.nih.gov/articles/PMC3772742/> |
| **NA (N1)** | D199G | Oseltamivir | Reduced | <https://pmc.ncbi.nlm.nih.gov/articles/PMC12270646/> |
| **NA (N1)** | D199Y | Oseltamivir | Reduced | <https://pmc.ncbi.nlm.nih.gov/articles/PMC12270646/> |
| **PA (Cap-Dependent Endonuclease)** | I38T | Baloxavir marboxil | 30-50 fold (H1N1); >50 fold (H3N2) | <https://www.nature.com/articles/s41598-018-27890-4> \| <https://pmc.ncbi.nlm.nih.gov/articles/PMC10786730/> |
| **PA (Cap-Dependent Endonuclease)** | I38M | Baloxavir marboxil | Reduced susceptibility | <https://www.nature.com/articles/s41598-018-27890-4> \| <https://pmc.ncbi.nlm.nih.gov/articles/PMC10786730/> |
| **PA (Cap-Dependent Endonuclease)** | I38F | Baloxavir marboxil | 24-fold (H5N1) | <https://pmc.ncbi.nlm.nih.gov/articles/PMC10786730/> |
| **PA (Cap-Dependent Endonuclease)** | I38N | Baloxavir marboxil | Reduced (less than I38T) | <https://wwwnc.cdc.gov/eid/article/31/5/24-1123_article> |
| **PA (Cap-Dependent Endonuclease)** | I38L | Baloxavir marboxil | 14.1-fold reduced | <https://journals.asm.org/doi/10.1128/aac.00009-22> \| <https://pmc.ncbi.nlm.nih.gov/articles/PMC9012239/> |
| **PA (Cap-Dependent Endonuclease)** | E199D | Baloxavir marboxil | 14.1-fold reduced | <https://journals.asm.org/doi/10.1128/aac.00009-22> \| <https://pmc.ncbi.nlm.nih.gov/articles/PMC9012239/> |
| **PA (Cap-Dependent Endonuclease)** | E23K | Baloxavir marboxil | 2-5 fold reduced | <https://www.sciencedirect.com/science/article/abs/pii/S0166354220302217> \| <https://www.nature.com/articles/s41598-018-27890-4> |
| **PA (Cap-Dependent Endonuclease)** | E23G | Baloxavir marboxil | Reduced susceptibility | <https://pmc.ncbi.nlm.nih.gov/articles/PMC10748225/> |
| **PA (Cap-Dependent Endonuclease)** | A37T | Baloxavir marboxil | Reduced susceptibility | <https://pmc.ncbi.nlm.nih.gov/articles/PMC10373543/> |
| **M2 (Ion Channel; Adamantane)** | S31N | Amantadine; Rimantadine | Near universal (>99% circulating) | <https://www.who.int/teams/global-influenza-programme/laboratory-network/quality-assurance/antiviral-susceptibility-influenza> |
| **M2 (Ion Channel; Adamantane)** | L26F | Amantadine; Rimantadine | Rare clinically | \| <https://pmc.ncbi.nlm.nih.gov/articles/PMC547263/> |
| **M2 (Ion Channel; Adamantane)** | V27A | Amantadine; Rimantadine | Predominant in H1N1 historically | <https://pmc.ncbi.nlm.nih.gov/articles/PMC547263/> |
| **M2 (Ion Channel; Adamantane)** | A30T | Amantadine; Rimantadine | Rare | <https://pmc.ncbi.nlm.nih.gov/articles/PMC547263/> |
| **M2 (Ion Channel; Adamantane)** | G34E | Amantadine; Rimantadine | Rare | <https://pmc.ncbi.nlm.nih.gov/articles/PMC547263/> |
| **M2 (Ion Channel; Adamantane)** | V27A/S31N | Amantadine; Rimantadine | Synergistic effects reduced | <https://pmc.ncbi.nlm.nih.gov/articles/PMC547263/> |
| **HA (Receptor Binding; Compensatory)** | K130N | Oseltamivir; Zanamivir (indirect) | 10-fold oseltamivir resistance | <https://www.nature.com/articles/s41467-025-66307-5> \| <https://pmc.ncbi.nlm.nih.gov/articles/PMC12749811/> |
| **HA (Receptor Binding; Compensatory)** | K130E | Oseltamivir; Zanamivir | Stronger resistance but high fitness cost | <https://pmc.ncbi.nlm.nih.gov/articles/PMC12749811/> |
| **HA (Receptor Binding; Compensatory)** | D225G | Compensatory (with NA mutations) | Rescues small-plaque phenotype of Y155H | <https://pmc.ncbi.nlm.nih.gov/articles/PMC3772742/> \| <https://academic.oup.com/jid/article/201/10/1517/992777> |
| **HA (Receptor Binding; Compensatory)** | D225N | Compensatory (with NA mutations) | Reduced drug susceptibility | <https://pmc.ncbi.nlm.nih.gov/articles/PMC3772742/> |
| **HA (Receptor Binding; Compensatory)** | N156K | Compensatory | Co-occurs with K130N | <https://www.nature.com/articles/s41467-025-66307-5> |
| **HA (Receptor Binding; Compensatory)** | A186T | Compensatory | Co-occurs with K130N | <https://www.nature.com/articles/s41467-025-66307-5> |
| **HA (Receptor Binding; Compensatory)** | Q189E | Compensatory | Receptor binding diversity | <https://pmc.ncbi.nlm.nih.gov/articles/PMC11118561/> |
| **HA (Receptor Binding; Compensatory)** | E224A | Compensatory | Clade-defining (5a.2a) | <https://pmc.ncbi.nlm.nih.gov/articles/PMC11118561/> |
| **HA (Receptor Binding; Compensatory)** | P137S | Compensatory | Clade-defining (5a.2a.1) | <https://pmc.ncbi.nlm.nih.gov/articles/PMC11118561/> |
| **HA (Receptor Binding; Compensatory)** | K142R | Compensatory | Clade-defining (5a.2a.1) | <https://pmc.ncbi.nlm.nih.gov/articles/PMC11118561/> |

**Supplementary table 3. Mutations compared to the reference H1N1 strain** (References used for comparison: *Hemagglutinin (HA) - Segment 4: NC_026433.1*

*Neuraminidase (NA) - Segment 6: NC_026434.1*

*Matrix (M1, M2) - Segment 7: NC_026431.1*

*Polymerase Acidic (PA) - Segment 3: NC_026437.1*).

| **Protein** | Location (aa) | Reference (2009 pandemic strain) | Consensus sequence from the sample on 18th of February |
| --- | --- | --- | --- |
| **HA** | 8 | L | M |
| **HA** | 13 | A | T |
| **HA** | 36 | V | L |
| **HA** | 71 | K | Q |
| **HA** | 91 | S | R |
| **HA** | 220 | S | T |
| **HA** | 233 | I | T |
| **HA** | 241 | E | A |
| **HA** | 267 | V | A |
| **HA** | 273 | A | T |
| **HA** | 276 | R | K |
| **HA** | 277 | N | D |
| **HA** | 300 | K | E |
| **HA** | 312 | I | V |
| **HA** | 325 | K | R |
| **HA** | 338 | I | V |
| **HA** | 516 | E | K |
| **HA** | 523 | E | D |
| **HA** | 527 | I | T |
| M2 | 21 | D | G |
| M2 | 23 | S | N |
| **NA** | 188 | I | T |
| **NA** | 222 | N | K |
| **NA** | 241 | V | I |
| **NA** | 248 | N | D |
| **NA** | 264 | V | A |
| **NA** | 314 | I | M |
| **NA** | 321 | I | V |
| **NA** | 369 | N | K |
| **NA** | 386 | N | K |
| **NA** | 389 | I | K |
| **NA** | 416 | D | N |
| **NA** | 432 | K | E |
| **NA** | 449 | N | D |
| **NA** | 452 | T | I |
| **NA** | 453 | V | M |
| PA | 5 | V | L |
| PA | 63 | V | I |
| PA | 100 | V | I |
| PA | 224 | P | S |
| PA | 225 | S | C |
| PA | 262 | R | K |
| PA | 321 | N | K |
| PA | 330 | I | V |
| PA | 335 | L | F |
| PA | 337 | A | T |
| PA | 362 | R | K |
| PA | 438 | I | V |
| PA | 505 | I | V |

**Supplementary table 3.** Single nucleotide variant analyses for resistance sites.

| **virus** | **gene** | **aa_pos** | **ref_aa** | **expected_mutations** | **total_reads** | **resistance**  **reads** | **resistance**  **percent** | **aa_counts** |
| --- | --- | --- | --- | --- | --- | --- | --- | --- |
| A(H1N1)pdm09 | HA | 130 | R | E/N | 1781 | 0 | 0.0 | K:2;M:3;R:1774;S:2 |
| A(H1N1)pdm09 | HA | 137 | T | S | 1679 | 0 | 0.0 | A:1679 |
| A(H1N1)pdm09 | HA | 142 | N | R | 1534 | 0 | 0.0 | N:1534 |
| A(H1N1)pdm09 | HA | 156 | A | K | 1728 | 0 | 0.0 | A:1727;V:1 |
| A(H1N1)pdm09 | HA | 186 | K | T | 0 | 0 | 0.0 | Not covered |
| A(H1N1)pdm09 | HA | 189 | E | E | 0 | 0 | 0.0 | Not covered |
| A(H1N1)pdm09 | HA | 224 | S | A | 11318 | 0 | 0.0 | I:6;N:8;S:11304 |
| A(H1N1)pdm09 | HA | 225 | K | G/N | 10702 | 2 | 0.019 | K:10700;N:2 |
| A(H1N1)pdm09 | NA | 114 | V | I | 1072 | 0 | 0.0 | V:1072 |
| A(H1N1)pdm09 | NA | 136 | Q | K | 1380 | 0 | 0.0 | Q:1380 |
| A(H1N1)pdm09 | NA | 155 | Y | H | 6014 | 0 | 0.0 | Y:6014 |
| A(H1N1)pdm09 | NA | 199 | D | G/Y | 8696 | 0 | 0.0 | D:8696 |
| A(H1N1)pdm09 | NA | 223 | I | R/T/V | 2989 | 1 | 0.033 | I:2987;L:1;V:1 |
| A(H1N1)pdm09 | NA | 247 | S | N | 7534 | 0 | 0.0 | I:7;S:7527 |
| A(H1N1)pdm09 | NA | 275 | H | Y | 9277 | 3 | 0.032 | H:9274;Y:3 |
| A(H1N1)pdm09 | NA | 295 | N | S | 8740 | 0 | 0.0 | D:2;N:8738 |
| A(H1N1)pdm09 | M2 | 26 | L | F | 38094 | 3 | 0.008 | F:3;L:38091 |
| A(H1N1)pdm09 | M2 | 27 | V | A | 38167 | 1 | 0.003 | A:1;F:13;I:6;V:38147 |
| A(H1N1)pdm09 | M2 | 30 | A | T | 38575 | 11 | 0.029 | A:38538;E:12;S:8;T:11;V:6 |
| A(H1N1)pdm09 | M2 | 31 | N | N | 43676 | 43663 | 99.97 | K:13;N:43663 |
| A(H1N1)pdm09 | M2 | 34 | G | E | 41441 | 1 | 0.002 | E:1;G:41435;R:1;V:3;W:1 |
| A(H1N1)pdm09 | PA | 23 | E | G/K | 3210 | 1 | 0.031 | *:4;E:3205;K:1 |
| A(H1N1)pdm09 | PA | 37 | A | T | 3361 | 1 | 0.03 | A:3352;E:3;P:2;S:3;T:1 |
| A(H1N1)pdm09 | PA | 38 | I | F/L/M/N/T | 5637 | 0 | 0.0 | I:5636;V:1 |
| A(H1N1)pdm09 | PA | 199 | E | D | 33522 | 0 | 0.0 | *:7;A:1;E:33500;K:13;Q:1 |

**Supplementary table 4.** Resistance mutations checked in the consensus sequence of RSV-A.

| **Position** | **Wild Type** | **Mutant** | **Target / Context** | **Source** | **PMID/DOI** |
| --- | --- | --- | --- | --- | --- |
| **429** | Arginine (R) | S (Serine) | Clesrovimab (Site IV). Critical binding residue. | Tang et al. (2019) Nature Communications | DOI: 10.1038/s41467-019-12137-1 |
| **432** | Isoleucine (I) | T (Threonine) | Site V/VI (historical mAbs). Reduced binding. | López et al. (1998) J. Virol | PMID: 9658147 |
| **433** | Lysine (K) | T (Threonine) | Site V/VI & Clesrovimab. Drastic reduction in binding. | López et al. (1998) J. Virol; | PMID: 9658147 |
| **436** | Serine (S) | F (Phenylalanine) | Site VI (historical mAbs). | López et al. (1998) J. Virol | PMID: 9658147 |
| **443** | Serine (S) | P (Proline) | Clesrovimab. Strong resistance mutation. | Tang et al. (2019) Nature Communications | DOI: 10.1038/s41467-019-12137-1 |
| **446** | Glycine (G) | E (Glutamine) | Strong resistance mutation. | Tang et al. (2019) Nature Communications | DOI: 10.1038/s41467-019-12137-1 |
| **447** | Valine (V) | A (Alanine) | Site IV/V. Historical escape mutant (C-terminal boundary of epitope). | López et al. (1998) J. Virol; Mousa et al. (2015) Curr Opin Virol | PMID: 9658147 (López); PMID: 25819327 (Mousa) |
